# Performance of serum apolipoprotein-A1 as a sentinel of Covid-19

**DOI:** 10.1101/2020.09.01.20186213

**Authors:** Thierry Poynard, Olivier Deckmyn, Marika Rudler, Valentina Peta, Yen Ngo, Mathieu Vautier, Sepideh Akhavan, Vincent Calvez, Clemence Franc, Jean Marie Castille, Fabienne Drane, Mehdi Sakka, Dominique Bonnefont-Rousselot, Jean Marc Lacorte, David Saadoun, Yves Allenbach, Olivier Benveniste, Frederique Gandjbakhch, Julien Mayaux, Olivier Lucidarme, Bruno Fautrel, Vlad Ratziu, Chantal Housset, Dominique Thabut, Patrice Cacoub

## Abstract

**Background:** Since 1920, a decrease in serum cholesterol has been identified as a marker of severe pneumonia. We have assessed the performance of serum apolipoprotein-A1, the main transporter of HDL-cholesterol, to identify the early spread of coronavirus disease 2019 (Covid-19) in the general population and its diagnostic performance for the Covid-19.

**Methods:** We compared the daily mean serum apolipoprotein-A1 during the first 30 weeks of 2020 in a population that is routinely followed for a risk of liver fibrosis risk in the USA (183,112 sera) and in France (18,316 sera) in relation to a local increase in confirmed cases, and in comparison to the same period in 2019 (respectively 234,881 and 26,056 sera). We prospectively assessed the sensitivity of this marker in an observational study of 136 consecutive hospitalized cases and retrospectively evaluated its specificity in 7,481 controls representing the general population.

**Results:** The mean serum apolipoprotein-A1 levels in these populations began decreasing in January 2020, compared to the same 30 weeks in 2019. This decrease was highly correlated to and in parallel with the daily increase in confirmed Covid-19 cases in the following 30 weeks, in both France and USA, including the June and mid-July recovery periods in France. Apolipoprotein-A1 at the 1.25 g/L cutoff had a sensitivity of 90.6% (95%CI84.2-95.1) and a specificity of 96.1% (95.7-96.6%) for the diagnosis of Covid-19. The area under the characteristics curve was 0.978 (0.957-0.988), and outperformed haptoglobin and liver function tests. The adjusted risk ratio for survival without transfer to intensive care unit was 5.61 (95%CI 1.02-31.0;P=0.04).

**Conclusion:** Apolipoprotein-A1 could be both a sentinel of the pandemic in existing routine surveillance of the general population with no new blood sample, as well as a candidate predictor of suspected Covid-19 in multivariate analysis in cases with a negative virological test. NCT01927133, CER-2020-14.

*Key Points:* 

*Question:* Does serum apolipoprotein-A1 decrease could be a very early biomarker of SARS-CoV-2 pandemic?

*Findings:* During the 30 weeks of 2020 we observed in two large cohorts of patients at risk of liver fibrosis a highly significant decrease of serum apolipoprotein-A1, not observed in previous years. This decrease was highly correlated to and in parallel with the daily increase in confirmed Covid-19 cases, including the recovery period.

*Meaning:* Apolipoprotein-A1 could be used as a sentinel of the pandemic in existing routine surveillance of the general population.

---

**Cover Letter**

Dear Editor

We submit to PlosMedicine an article entitled “**Performance of serum apolipoprotein-A1 as a sentinel of Covid-19**”.

I share my professional life with apolipoprotein-A1 (ApoA1) since 1982, with a prospective cohort of 975 heavy French drinkers, followed by several other cohorts looking for fibrosis biomarkers.

In February 2020, serendipity scenario.

Our Hepatology department of Pitié-Salpêtrière, was transformed in a Covid-19 center and I rapidly observed in these severe patients a specific profile of Apo-A1 decrease and Haptoglobin increase, never observed before.

The night after, I realize that we have online more than 1 million of ApoA1 and Haptoglobin in the cohorts of patients followed by FibroTest for a risk of liver fibrosis. Therefore, we compared daily the ongoing year 2020 to previous years “Covid-Free”. Indeed, there was a highly significant decrease in ApoA1 parallel (2 weeks before) to the confirmed Covid-19 cases.

The most original result was that this decrease started few weeks before the incidence of confirmed cases, suggesting that ApoA1 detected infected cases. Furthermore, the liver function biomarkers as well as the Haptoglobin did not change so early.

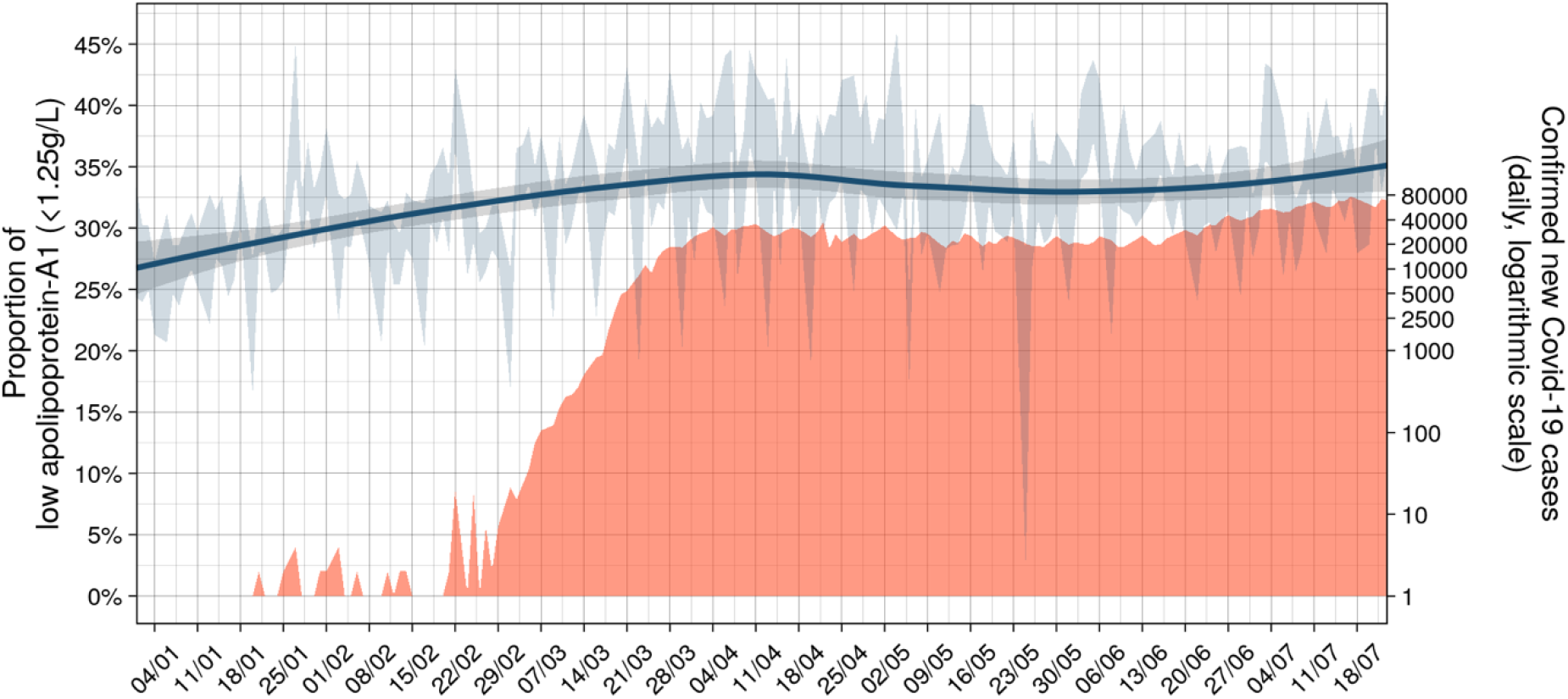

We observed the normalization of apolipoprotein-A1 in the sera of the French cohort in June, in parallel with the decrease of daily confirmed Covid-19 cases, as well as the plateau in the US cohort.

Apolipoprotein-A1 could be a very simple early marker of the Covid-19 pandemic, as suggested in two ongoing surveillance databases, in France and USA with high sensitivity and specificity. It can be done immediately and with low cost in routine surveillance already dosing apolipoprotein-A1, as well as in patients with a clinical suspicion of Covid-19 but with a negative virologic test.

We do think that this article will help clinicians immediately, in the phase of a possible second wave, and epidemiologists for estimating the prevalence of infected subjects in general population, using other surveillance cohorts. Furthermore, our results suggest a potential intestinal transmission without liver signals.

This manuscript has not been presented or is not submitted elsewhere. None of the material concerning the large surveillance databases and the prospective observational cohort has been published or is under consideration for publication elsewhere.

According to the context of the pandemic, for assessing the specificity we used controls previously published in large studies of liver diseases. This permitted to have a wide spectrum of false positive risk. These measurements have been all performed on fresh serum collected prospectively and analyzed in the biochemistry unit of Pitié-Salpêtrière Hospital, Paris, France, with the same biochemical methods as for the prospective Covid-19 cases.

The prospective observational study in Covid-19 patients was approved by CER-Sorbonne University IRB, N° 2020-CER-2020-14, with a signed informed consent. Clinical investigation was conducted according to the principles of the Declaration of Helsinki. All authors had access to the study data and reviewed and approved the final manuscript. The components of Fibrotest and Nash-FibroTest were measured in routine in all patients with a risk of liver disease in France. The different databases integrated were used on sera anonymously. All the retrospective database analyses previously published either non-interventional study, without supplementary blood sample, were exempt from institutional review board (IRB) review (ethical committee of ‘Comité de Protection des Personnes of Paris, Ile-de-France’ FIBROFRANCE project. CPP-IDF-VI, 10-1996-DR-964, DR-2012-222 and USA-NCT01927133).

This project was funded by the European Grant EIT health ProCoP 20879, Patrice Cacoub being the awarded author.

All the coauthors have reviewed this manuscript.

The data that support the findings of this study are fully available from the corresponding author, upon request.

I have a potential conflict of interest as the inventor of FibroTest, and founder of BioPredictive a spinoff company of Sorbonne University. The patent belongs to the French public organization Assistance Publique Hôpitaux de Paris.

Kind regards

Thierry Poynard

## Introduction

There is an urgent need to detect people in the general population who are at risk of being admitted to the hospital for Covid-19. Although viral nucleic acid testing and chest computed tomography are standard methods for diagnosing Covid-19 in patients with symptoms, these are time consuming. A review reported that there are only three models for predicting hospital admission in the healthy general population, and these are limited by a high risk of bias, and by using proxy outcomes.^1^

In the meantime, several early detection tests of cytokine burst could be immediately available. In 1920, Harold A. Kipp found that a decrease in serum cholesterol was a marker of severe pneumonia.^2^ One hundred years later, a meta-analysis confirmed that levels of high-density lipoprotein cholesterol (HDL) and a level below the median of its transporter, apolipoprotein-A1, was associated with a two-fold increase in mortality in patients with severe sepsis (**supplementaryFig1,supplementaryTable1**).^3^ Its decrease seems to occur earlier than the increase in haptoglobin, another marker of sepsis.^4,5^ Furthermore, unlike haptoglobin, which is synthesized mainly by the liver, apolipoprotein-A1 is synthesized by both the liver and the intestine.^5^ This hypolipidemia begins in Covid-19 patients with mild symptoms.^6,7^ Therefore, in the general population, apolipoprotein-A1 might be a sensitive marker of infection in asymptomatic subjects, in those without the pulmonary symptoms of standard Covid-19, or in patients with an intestinal route of infection (**supplementaryFile1**).^8,9^

## Methods

### Ethics

The prospective observational study in Covid-19 patients was approved by CER-Sorbonne University IRB,CER-2020-14, with a signed informed consent. All of the previously published patient analyses from retrospective databases were non-interventional studies, without supplementary blood samples, and were exempt from a review of the IRB (NCT01927133,**supplementaryFile2**). The investigation was performed according to the principles of the Declaration of Helsinki. All authors had access to the study data and reviewed and approved the final manuscript.

### Decrease of apolipoprotein-A1

We used three cohorts of sera from subjects at risk of liver fibrosis followed by FibroTest (FibroSure in USA,**supplementaryFile2**),^10^ a large private-laboratories US cohort (“US-cohort”), a cohort with an intermediate risk of Covid-19 “(French-cohort”) were patients being followed in academic and private-laboratories, and a high-risk cohort which included patients at the Pitie-Salpetriere hospital Paris, France (“APHP-PSL”). The core temporal analysis compared the first 30 weeks of consecutive anonymous sera 2020 vs. the sera 2019, from these three routinely followed cohorts (**supplementaryFile2**, **supplementaryFig2C**).

### Confounding factors

Decrease in apolipoprotein-A1 may be due to direct liver toxicity from SARS-CoV-2,^11^ but also to drug-induced liver disease (DILI) caused by medications (**supplementaryFile2**, **supplementaryTable2**). We analyzed the kinetics of alpha2-macroglobulin (A2M) a specific marker of liver fibrosis,^10^ and of haptoglobin, a sensitive biomarker of severe acute phase, as well in the US-cohort of all the other components, according to non-alcoholic fatty liver disease (NAFLD) sera, or chronic hepatitis C (HCV) sera(**supplementaryFile2**).

### Daily apolipoprotein-A1 and spread of Covid-19

The number of confirmed Covid-19 cases in France and in the USA was assessed according to published data from the European Centre for Disease Prevention and Control (https://ourworldindata.org/coronavirus-data) (**supplementaryFig2A**, **supplementaryFig2B**).

### Sensitivity and prognostic value

Sensitivity was assessed in a prospective study of Covid-19 patients hospitalized in APHP-PSL, described in **supplementaryFile2**. The primary endpoint was the survival without transfer in ICU at 28 days, adjusted on age, gender, haptoglobin, and liver tests. (**supplementaryTable1**, **supplementaryTable2**,**supplementaryFile1**.

### Specificity

We collected prospective data on apolipoprotein-A1 from and since the first cohort of alcoholic liver disease in 1982, and from the FibroFrance cohort (**supplementaryFile2**). More recently, we collected six cohorts, which allowed us to retrospectively validate the specificity in a large group of subjects (**supplementaryFile2**).^12-16^ The measurements were all performed on fresh prospectively collected serum and analyzed in the biochemistry unit of the APHP-PSL hospital, with the same methods as the Covid-19 cases (**supplementaryTable3**). The core control population was a group of healthy volunteers that was representative of the French population.^16^ Forty-three patients without suspected Covid-19 were excluded (**supplementaryTable4**). In order to identify a profile of patients with a possible intestinal route of infection, we compared the subsets of patients with or without diarrhea.

Apolipoprotein-A1, haptoglobin, A2M, gammaglutamyl transpeptidase (GGT), alanine aminotransferase (ALT) were assessed (**supplementaryTable2**,**supplementaryFile1**), following BioPredictive (Paris, France) analytical recommendations.^17^ The virological methods for the diagnosis used to diagnose SARS-CoV2 in respiratory samples, were detailed in **supplementaryFile2**.

## Statistical methods

The first primary endpoint was to demonstrate a significant decrease of the mean daily value of apolipoprotein-A1 in the 30 weeks of the year 2020, *vs*. those of the year 2019. We defined a significant decrease of apolipoprotein-A1 as below 1.25 g/L, the optimal cutoff defined by the highest Youden index (sensitivity specificity −1) in the severe patients. The daily proportion of sera below this cutoff defined a significant risk of Covid-19. The second analysis was to exclude confounding factors detailed in **supplementaryFile1**. We compared the linear regression curves with 95% confidence intervals (95%CI) between the mean daily levels of all markers, and the daily proportion of low apolipoprotein-A1, between the same 30 weeks periods per year, by the *F*-test. A significant difference was defined as P<0.001. The third primary endpoint was to assess the association between the spread of Covid-19 and the decrease in apolipoprotein-A1. The diagnostic performance of biomarkers was assessed using non-parametric AUROCs. The prognostic values were assessed by survival curves, using Kaplan-Meir method, compared by Logrank test (cutoff being the median of this context of use) and adjusted by Cox model. The repeated sera assessments were compared by repeated ANOVA and Tukey-Kramer multiple-comparison test. R and NCSS-2020 were used as statistical software.

## Results

### Decrease of apolipoprotein-A1

The mean daily levels of apolipoprotein-A1 decreased globally (**fig1A**) and in the three cohorts (**fig1B**; all P<0.001), during the first 30 weeks of 2020. This was already highly significant in January 2020 in the US-cohort (lower panel), compared to first 30 weeks of 2019 and 2018. The mean daily proportion of sera with low apolipoprotein-A1 was 32.4%(95%CI 32.4-32.5) on 30 weeks, that is 5.1% higher *vs*. 26.8%(27.2-27.4;P<0.001) in 2019 (**fig1C**). There was no significant difference between the years 2019 and 2018 for the daily mean globally (**fig1A**), or by cohort (**fig1B**). The proportion of low apolipoprotein-A1, in 2018 was 26.5%(26.3-26.6), that is a small 0.8% difference vs. the year 2019 (**fig1C**).

**Fig 1.**
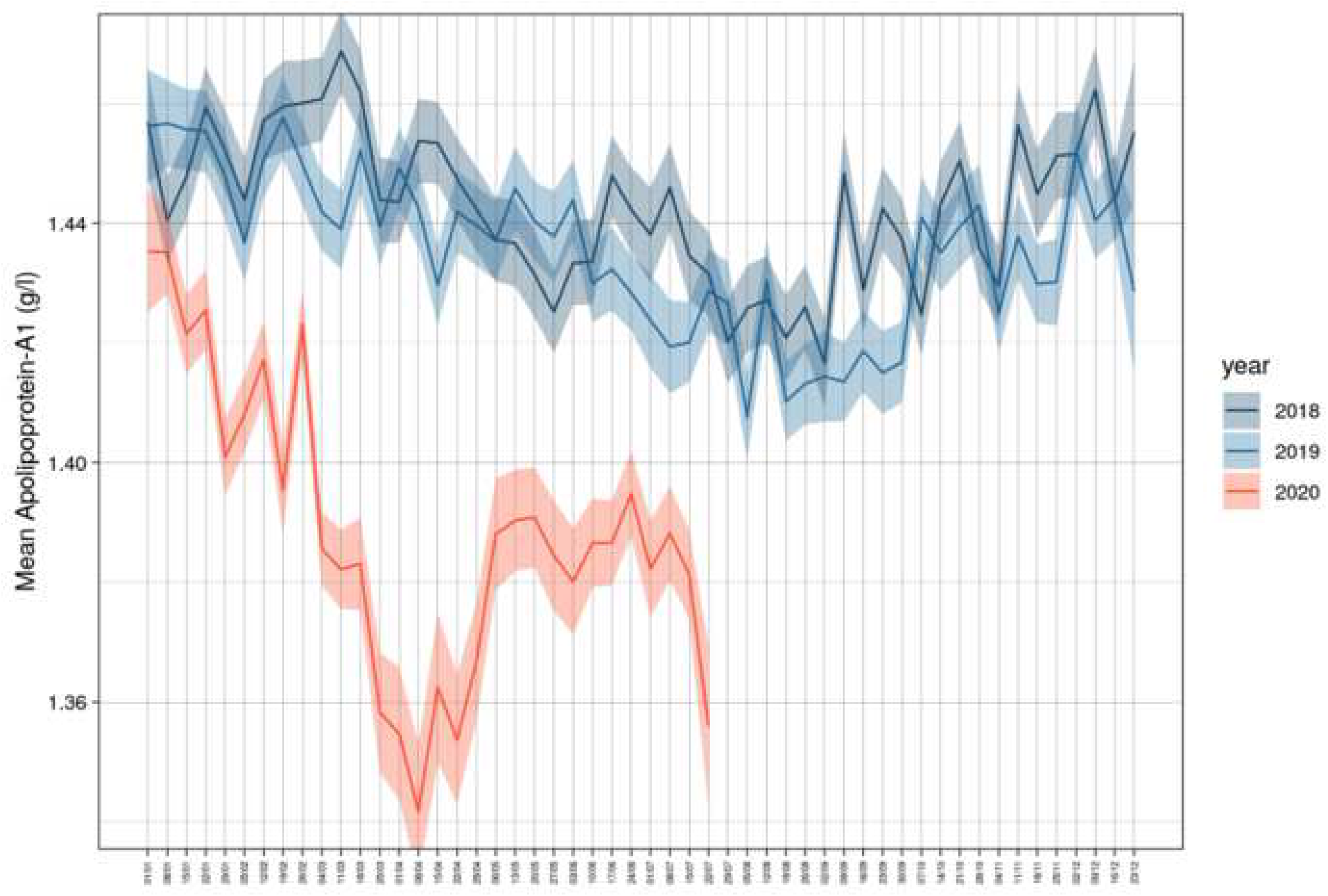
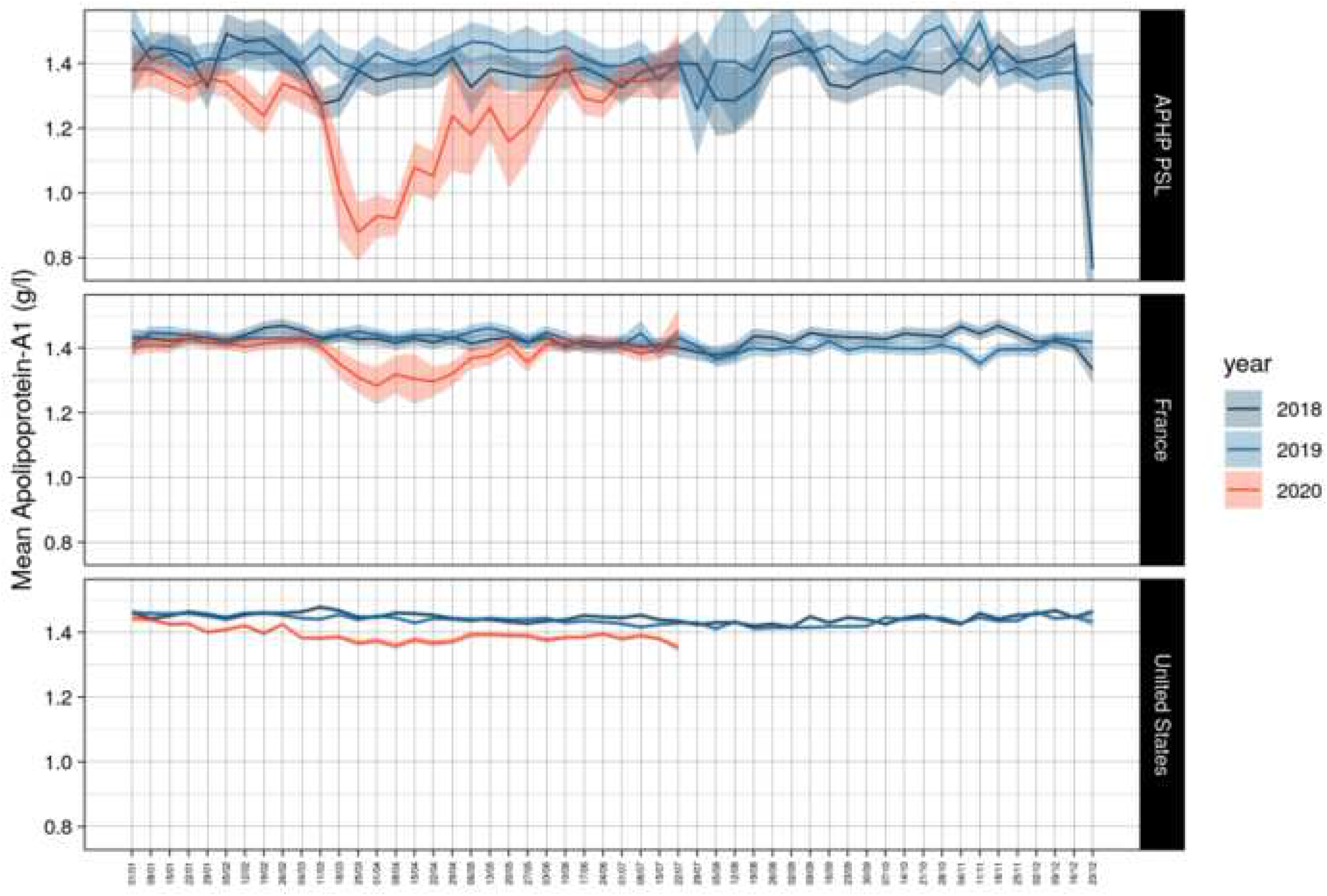
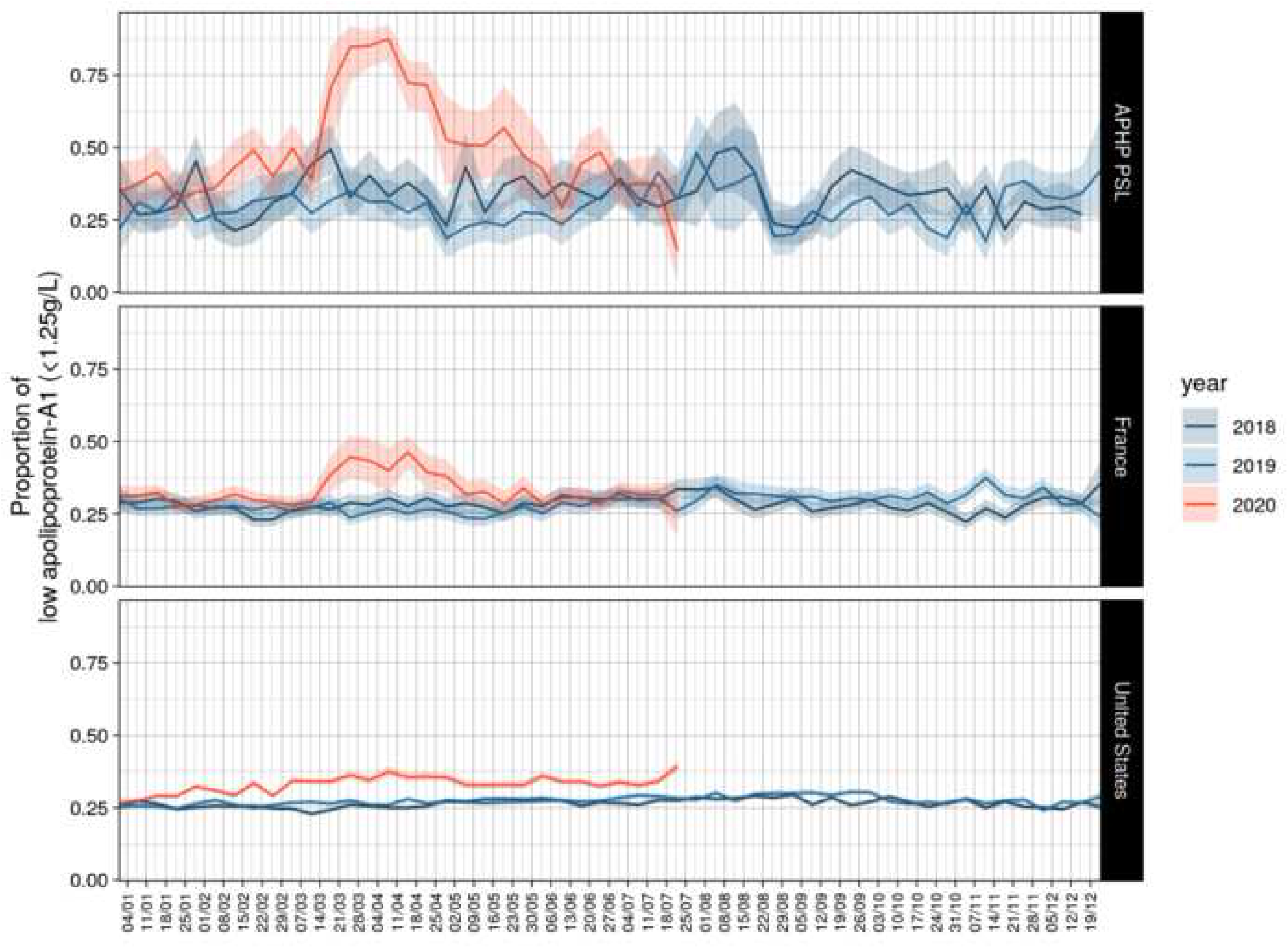
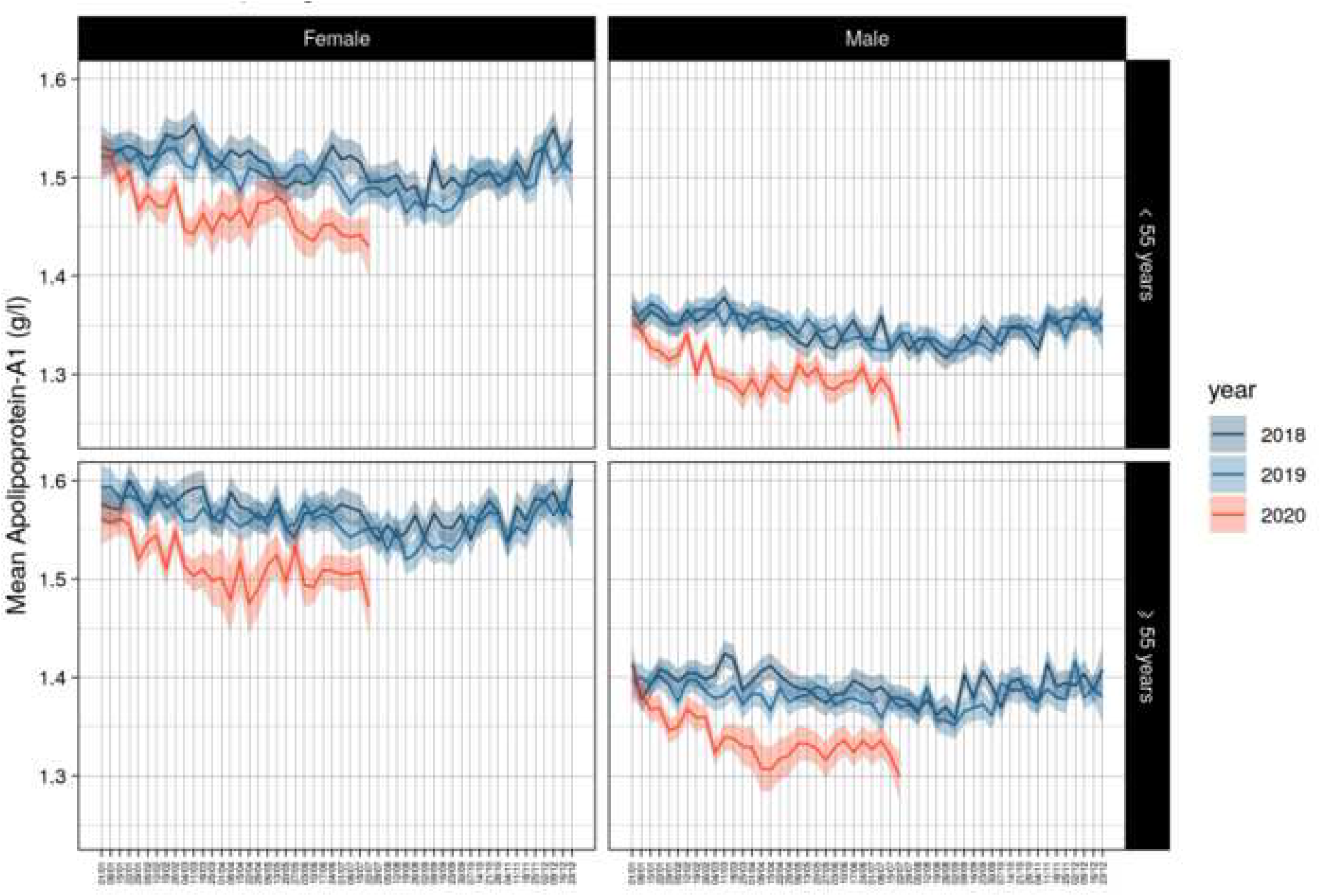
Decrease of apolipoprotein-A1 value in the first 30 weeks of 2020. **1A Global decrease of apolipoprotein-A1 among the sera of patients with risk of liver fibrosis in the first 30 weeks of 2020 in USA and France compared to the two previous years** (P<0.001). **1B Quantitative decrease of apolipoprotein-A1 by cohort**. Apolipoprotein-A1 decreased (P<0.001) in the three cohorts, starting early in January 2020 (red line with 95% confidence interval) in the US cohort (lower panel). **1C Proportion of serum with low apolipoprotein-A1 by cohorts**. Low apolipoprotein-A1 was defined as below 1.25 g/L. Details in **supplementaryFile2**. **1D Decrease of apolipoprotein-A1 by gender and age in US cohort**. The same significant kinetics were observed, P<0.001 between 2020 and previous years. **1E Number of confirmed Covid-19 cases per day and proportion of low (<1.25g/L) apolipoprotein-A1 in the US cohort during the first 30 weeks of 2020**. The red graph is the number of confirmed cases per day in logarithmic scale. The black line is the daily mean proportion of low apolipoprotein-A1 (<1.25g/L; blue line;95%CI in grey).

### Confounding factors

The same significant kinetics in apolipoprotein-A1 levels were observed after stratification of the regression curves for gender and age in the three cohorts (**supplementaryFig3**). The US-cohort was the only sample that had the necessary power to compare these two factors together between 2020 and 2019 and 2018 (**fig1D**), and in the subset of patients with HCV (**supplementaryFig3C**). Apolipoprotein-A1 decrease was similar during Covid-19 spread versus 2019 and 2018 in the US-cohort(**supplementaryFile2**, **supplementaryFig3**).

The kinetics of apolipoprotein-A1 were not associated with those of haptoglobin (**supplementaryFig4**).

For A2M, there was in the US-cohort only a significant lower mean serum value, when compared to 2019 and even more compared to 2018 (**supplementaryFig5A**), and detailed in **supplementaryFig2**, **supplementaryFig5**), and highest in HCV *vs*. NAFLD sera. After adjustment on age and gender there was no significant decrease of A2M between 2020 and 2019 (**supplementaryFig5**).

The other significant differences were, in the French-cohorts, GGT increased during the pandemic peak and returned to previous years’ value thereafter (**supplementaryFig6**), and an increase in ALT in April 2020, in the US-cohort (**supplementaryFig7**).

No significant changes were observed for total cholesterol (**supplementaryFig8**), triglycerides (**supplementaryFig9**), fasting glucose (**supplementaryFig10**), weight (**supplementaryFig11**) and height (**supplementaryFig12**).

### Temporal associations between Covid-19 cases and apolipoprotein-A1

The daily mean number of confirmed Covid-19 cases paralleled about 10 days after the daily proportion of low apolipoprotein-A1 in sera. In USA, the first ten Covid-cases were declared mid-January 2020, when the proportion of low apolipoprotein-A1 had already increase by several percent (**fig1E**). The first peak of cases (n=50,000) was reached mid-April (**supplementaryFig2B**), and the first peak of low apolipoprotein-A1 (37.0%) on April 7^th^ (**fig1E**, **supplementaryFile2**).

In France, the first ten Covid-cases were declared first week of March 2020 (**supplementaryFig2A**), when the proportion of low apolipoprotein-A1 started to increase by several percent (**fig1C**). The first peak of cases (n=7,000) was reached mid-April (**supplementaryFig2A**), as well as the first peak of low apolipoprotein-A1 (40.0%) on April 14^th^ (**fig1C**, **supplementaryFile2**).

### Diagnostic performance of apolipoprotein-A1

A total of 136 consecutive patients with severe Covid-19, but who did not require ICU were include. Their characteristics and were similar to those published in such severity profiles (Table 2,**supplementaryFile3**).

**Table 1.** Characteristics of the sera included in US and French-cohorts

| Characteristics | 2020 | 2019 | P-Value |
| --- | --- | --- | --- |
| **US-cohort** | **N=183,112** | **N=234,881** |  |
| Median age [IQR] | 54.1 [40.4-63.3] | 55.0 [41.7-63.3] | <0.001 |
| Age category n (%) |  |  | <0.001 |
| < 50 year | 75,782(41.4) | 90,853 (38.5) |  |
| 50 to <70 year | 88,597 (48.4) | 121,757 (51.8) |  |
| >= 70 year | 18,733 (10.2) | 22,771 (9.7) |  |
| Male sex (%) | 103,672 (56.6) | 131,056 (55.8) | <0.001 |
| Apolipoprotein-A1<1.25 g/L (%) | 59,419 (32.4) | 62,898 (26.8) | <0.001 |
| **Median laboratory (IQR)** |  |  |  |
| Apolipoprotein-A1 g/L | 1.37 [1.19-1.58] | 1.41 [1.23-1.68] | <0.001 |
| Haptoglobin g/L | 1.23 [0.82-1.68] | 1.23 [0.82-1.69] | 0.72 |
| Alpha-2-macroglobulin g/L | 2.16 [1.65-2.93] | 3.07 [1.70-3.07] | <0.001 |
| GGT IU/L | 36 [21-77] | 38 [21-79] | 0.87 |
| ALT IU/L<sup>1</sup> | 38 [22-70] | 38 [23-68] | <0.001 |
| Total bilirubin micromol/L | 6.84 [5.13-10.26] | 6.84 [5.13-10.26] | 0.01 |
| Fibrosis stage by FibroTest (%) |  |  | <0.001 |
| F0 | 94,170 (51.4) | 116,666 (49.7) |  |
| F1 | 36,738 (20.1) | 47,123 (20.1) |  |
| F2 | 13,330 (7.3) | 17,834 (7.6) |  |
| F3 | 17,914 (9.8) | 24,514 (10.4) |  |
| F4 | 19,648 (10.7) | 27,030 (11.5) |  |
| Non interpretable | 1,312 (0.7) | 1,714 (0.7) |  |
| NAFLD (surveillance by NASH-FibroTest)<sup>2</sup> | N=37,298 (20.2) | N=42,271 (18.0) | <0.001 |
| Weight (Kg) | 88.9 (74.8-105.2) | 88.5 (74.8-104.3) | 0.003 |
| Aspartate amino transferase (AST) | 34 (25-52) | 34 (25-52) | 0.55 |
| Total cholesterol | 4.53 (3.80-5.33) | 4.53 (3.80-5.28) | 0.001 |
| Triglycerides | 1.57 (1.13-2.24) | 1.55 (1.11-2.21) | 0.02 |
| Fasting glucose | 5.67 (5.06-6.89) | 5.61 (5.06-6.78) | <0.001 |
| Table 3 continued |  |  |  |
| **French-cohort** | **N= 18,316** | **N=26,056** |  |
| Median age [IQR] | 51.5 (41.2-64.4) | 52.6 (39.7-63.1) | <0.001 |
| Age category n (%) |  |  | <0.001 |
| < 50 year | 7,514 (40.9) | 11,517 (44.2) |  |
| 50 to <70 year | 8,187 (44.4) | 11,340 (43.5) |  |
| >= 70 year | 2,615 (14.3) | 3,199 (14.3) |  |
| Male sex (%) | 10,619 (58.0) | 14,908 (57.2) | <0.001 |
| Apolipoprotein-A1<1.25 g/L (%) | 5,694 (31.1) | 7,057 (27.1) | <0.001 |
| **Median laboratory (IQR)** |  |  |  |
| Apolipoprotein-A1 g/L | 1.38 (1.20-1.59) | 1.40 (1.23-1.61) | <0.001 |
| Haptoglobin g/L | 1.18 (0.79-1.62) | 1.15 (0.78-1.57) | <0.001 |
| Alpha-2-macroglobulin g/L | 1.96 (1.52-2.61) | 2.01 (1.56-2.69) | <0.001 |
| GGT IU/L | 36 (21-79) | 34 (20-73) | <0.001 |
| ALT IU/L | 31 (21-50) | 31 (21-50) | 0.42 |
| Total bilirubin micromol/L | 8.7 (6-12) | 8.7 (6-12) | 0.20 |
| Fibrosis stage by FibroTest (%) |  |  | <0.001 |
| F0 | 9,437 (51.5) | 13,671 (52.5) |  |
| F1 | 4,080 (22.3) | 5,708 (21.9) |  |
| F2 | 1,328 (7.3) | 1,877 (7.2) |  |
| F3 | 1,664 (9.1) | 2,263 (8.7) |  |
| F4 | 1,655 (9.0) | 2,336 (9.0) |  |
| Non interpretable | 190 (0.8) | 201 (0.8) |  |
| NAFLD (surveillance by NASH-FibroTest)<sup>2</sup> | 4152 (19.2) | 6,110 (20.4) | <0.001 |
| Weight | 83 (71-97) | 83 (71-97) | 0.41 |
| Aspartate amino transferase (AST) | 30 (23-42) | 30 (23-41) | 0.47 |
| Total cholesterol | 4.79 (4.00-5.55) | 4.75 (4.00-5.62) | 0.30 |
| Triglycerides | 1.37 (1.00-1.96) | 1.40 (1.00-2.00) | 0.02 |
| Fasting glucose | 5.76 (5.15-6.98) | 5.72 (5.17-6.90) | 0.87 |

<sup>1</sup> Not normal distribution, the means of ALT were in 2020 43.4 IU/L vs 42.7 IU/L in 2019
<sup>2</sup> NASH-FibroTest included more components than FibroTest: weight, aspartate aminotransferase, total cholesterol, triglycerides, and fasting glucose

**Table 2.** Characteristics of Covid-19 patients of the prospective study.

| Characteristics | COVID-19 diagnostic |  | P value | Total |
| --- | --- | --- | --- | --- |
|  | PCR positive | PCR negative<br>Adjudicated |  |  |
| Number n (%) | 117 (100%) | 19 (100%) |  | 136 |
| Median age (IQR) year | 72 (57-82) | 65.1 (49-74) | 0.15 | 72 (59-83) |
| Age category |  |  | 0.16 |  |
| <50 year | 14 (12.0) | 5 (26.4) |  | 19 (14.0) |
| 50 to <70 year | 37 (31.6) | 7 (36.8) |  | 44 (32.3) |
| >=70 year | 66 (56.4) | 7 (36.8) |  | 73 (53.7) |
| Male sex | 66 (56.4) | 12 (63.2) | 0.58 | 78 (57.4) |
| Geographic origin |  |  | 0.53 |  |
| Caucasian | 67 (57.3) | 14 (73.7) |  | 81 (59.6) |
| Subsaharan | 17 (14.5) | 1 (5.3) |  | 18 (13.2) |
| North African, Middle East | 20 (17.1) | 2 (10.5) |  | 22 (16.9) |
| Asian | 13 (11.1) | 2 (10.5) |  | 15 (11.0) |
| Oxygen-support category |  |  | 0.75 |  |
| Invasive oxygen support | 0 | 0 |  | 0 (0-0) |
| Noninvasive oxygen support | 82 (70.1) | 14 (73.7) |  | 96 (70.6) |
| None | 35 (29.9) | 5 (26.3) |  | 40 (29.4) |
| Coexisting conditions |  |  |  |  |
| Hypertension | 65 (55.6) | 9 (47.4) | 0.51 | 74 (54.4) |
| Diabetes | 32 (27.4) | 3 (15.8) | 0.29 | 35 (25.7) |
| Hyperlipidemia | 32 (27.4) | 7 (36.8) | 0.40 | 39 (28.7) |
| Liver disease | 11 (9.4) | 4 (21.1) | 0.13 | 15 (11.03) |
| Pulmonary disease | 13 (11.6) | 1 (5.6) | 0.44 | 14 (10.8) |
| Severe disease associated | 64 (55.2) | 11(57.9) | 0.82 | 75 (55.6) |
| Tobacco (ongoing or stopped) | 35 (29.9) | 8 (42.1.3) | 0.29 | 43 (31.6) |
| Alcohol consumption | 15 (12.9) | 5 (26.3) | 0.13 | 20 (14.8) |
| Initial presentation |  |  |  |  |
| Anosmia (13 missing) | 12 (11.1) | 3 (20.0) | 0.32 | 15 (12.2) |
| Ageusia (13 missing) | 16 (14.8) | 3 (20.0) | 0.60 | 19 (15.5) |
| Headache (13 missing) | 10 (9.3) | 4 (26.7) | 0.05 | 14 (11.4) |
| Dyspnea (12 missing) | 59 (54.1) | 8 (53.3) | 0.95 | 67 (54.0) |
| Wheezing (12 missing) | 3 (2.75) | 0 (0.00) | 0.52 | 3 (2.42) |
| Cough without spitting (12 missing) | 47 (43.1) | 6 (40.0) | 0.82 | 53 (42.7) |
| Cough with spitting (12 missing) | 18 (16.5) | 2 (13.3) | 0.75 | 20 (16.1) |
| Fatigue | 61 (56.5) | 8 (53.3) | 0.82 | 69 (59.1) |
| Time clinic-inclusion (days) | 8 (5-13.8) | 11 (9-16.5) | 0.13 | 9 (5-14) |
| Followup (days) | 38 (27-53) | 28 (26-36) | 0.05 | 32 (27-49) |
| BMI (Kg/m<sup>2</sup>) | 25 (23-29) | 24 (22-28) | 0.56 | 25 (23-28) |
| Apolipoprotein <= 1.25g/L | 104 (88.9) | 19 (100) | 0.13 | 123 (90.4) |
| Median laboratory (IQR) |  |  |  |  |
| Apolipoprotein-A1 g/liter | 0.85 (0.73-1.06) | 0.74 (0.61-0.87) | 0.06 | 0.84 (0.70-1.03) |
| Haptoglobin g/liter | 3.16 (2.20-4.19) | 3.18 (1.59-3.86) | 0.59 | 3.16 (2.22-4.08) |
| Alpha-2 macroglobulin g/liter | 1.49 (1.24-2.06) | 1.46 (1.12-2.01) | 0.41 | 1.49 (1.22-2.05) |
| GGT IU per liter | 47 (28-114) | 58 (32-102) | 0.27 | 49 (30-114) |
| ALT IU per liter | 29 (20-47) | 40 (15-57) | 0.80 | 31 (20-51) |
| Total bilirubin micromol/L | 7 (6-8) | 9 (8-13) | 0.01 | 8 (5-12) |
| Platelets 10<sup>6</sup> per ml | 211 (162-278) | 279 (194-330) | 0.06 | 221 (164-287) |
| Creatinine µmol per liter | 78 (62-103) | 78 (70-120) | 0.97 | 78 (63-103) |
| Fasting glucose | 6 (5.4-8.0) | 6.6 (5.2-7.9) | 0.38 | 6.1 (5.4-7.9) |
| Albumin g/L (18 missing) | 31(27-33) | 29 (27-39) | 0.89 | 30 (27-33) |
| Procalcitonin (17 missing) | 0.13 (0.08-0.27) | 0.14 (0.09-0.23) | 0.98 | 0.13 (0.09-0.27) |
| C-reactive protein (13 missing) | 56 (20.3-95.6) | 101 (15.4-252.9) | 0.21 | 59 (20.3-102.3) |
| InterLeukin-6 (38 missing) | 31 (13.5-55.2) | 29 (8.05-119) | 0.96 | 31 (12.3-55.5) |
| CPK (18 missing) | 101 (51.5-243.3) | 47.5 (37.5-127) | 0.05 | 94 (45.8-233) |
| AST (11 missing) | 44 (30-60) | 34 (25-42) | 0.04 | 41 (30-59) |
| LDH (16 missing) | 347 (278-419) | 353 (289-440) | 0.68 | 347 (278-420) |
| D-dimer (24 missing) | 1050 (550-2030) | 1570 (1140-3980) | 0.09 | 1125 (570-2158) |
| Troponin (14 missing) | 16.4 (9.85-34.63) | 17.2 (9.56-36.5) | 0.99 | 16.4 (9.85-34.63) |
| Prothrombin time (16 missing) | 92 (83-100) | 82 (39-89) | 0.005 | 90 (81-100) |
| White cells |  |  |  |  |
| Neutrophil missing (14 missing) | 4185 (2773-6145) | 5945 (3963-7550) | 0.05 | 4420 (2928-6200) |
| Eosinophil (12 missing) | 0 (0-30) | 10 (0-150) | 0.14 | 0.05 (0-30) |
| Lymphocyte (14 missing) | 965 (670-1235) | 1150 (798-1700) | 0.25 | 975 (685-1295) |
| Hemoglobin g/L (12 missing) | 12 (11.2-13.6) | 12 (10.5-14.7) | 0.64 | 12 (11.1-13.8) |
| Treatment at risk of DILI |  |  |  |  |
| Paracetamol oral (2-4 g/day) | 43 (36.8) | 5 (26.3) | 0.38 | 48 (35.3) |
| Antibiotics | 91 (77.8) | 15 (78.9) | 0.91 | 106 (77.9) |
| None | 26 (22.2) | 4 (21.1) |  | 30 (22.1) |
| Without clavulinate | 49 (41.9) | 5 (26.3) |  | 54 (39.7) |
| With clavulinate | 68 (58.1) | 14 (73.7) |  | 82 (60.3) |
| Steroids | 12 (10.3) | 1 (5.3) | 0.49 | 13 (9.56) |
| Hydroxy chloroquine | 47 (40.1) | 6 (31.6) | 0.48 | 53 (39.0) |
| No Intensive care no death | 99 (84.6) | 17 (89.5) | 0.23 | 116 (85.3) |
| Transfer intensive care unit | 18 (15.4) | 2 (10.5) | 0.58 | 20 (14.7%) |
| Death (4 in intensive care unit) | 15 (12.8) | 1(5.3) | 0.34 | 16 (11.8) |

The characteristics of patients included for the assessment of specificity are presented in **supplementaryTable3**, the differences between characteristics of the subsets in **supplementaryTable3**, in **fig2A** for the median value of apolipoprotein-A1 and in **fig2B** for haptoglobin at inclusion.

**Fig 2.**
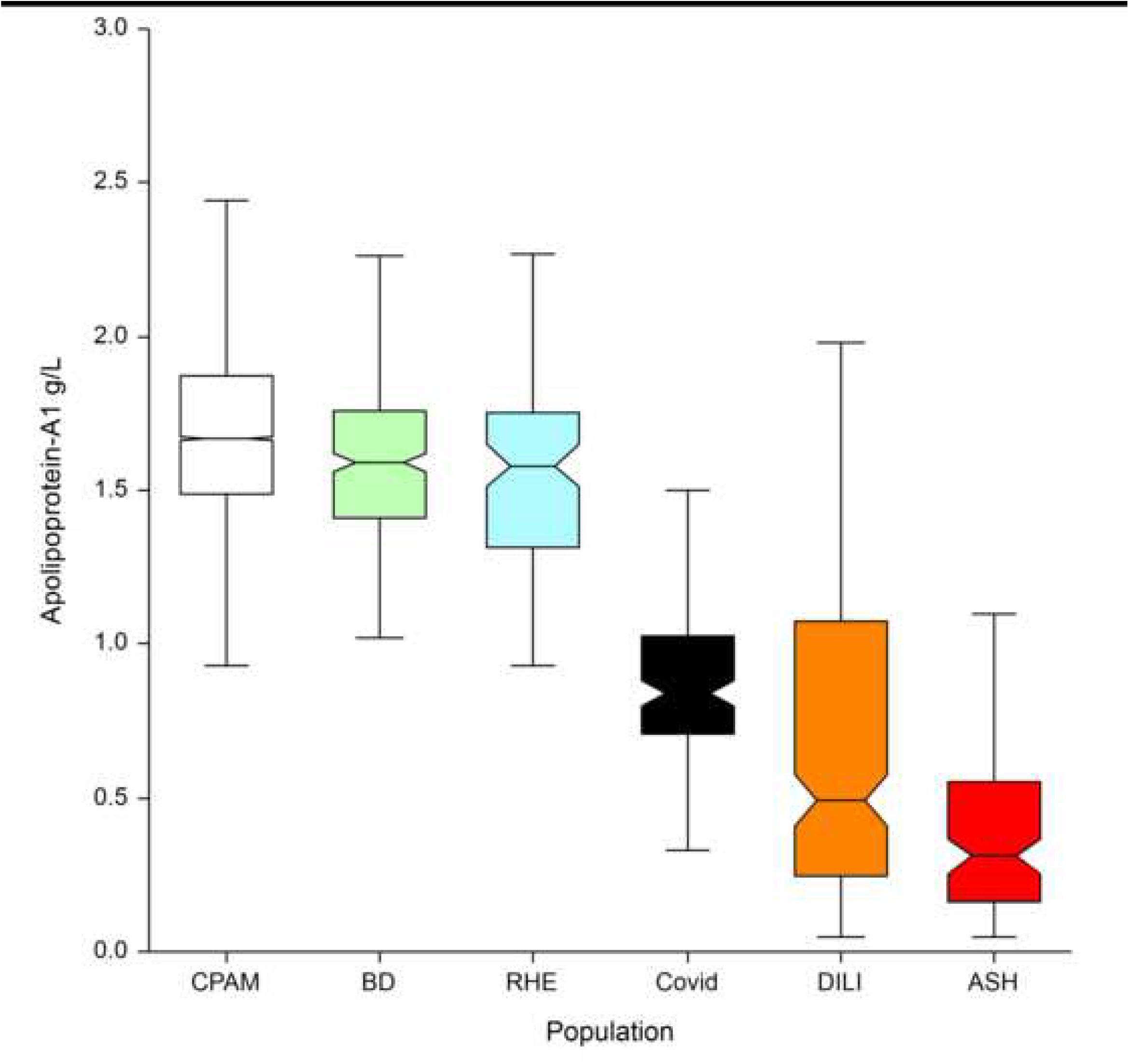
Performances of apolipoprotein-A1 and haptoglobin for the diagnostic and prognostic of Covid-19 patients. Details in supplementaryFile2. **Fig 2A**. **Apolipoprotein-A1 median with IQR between the 6 populations**

**Fig 2B.**
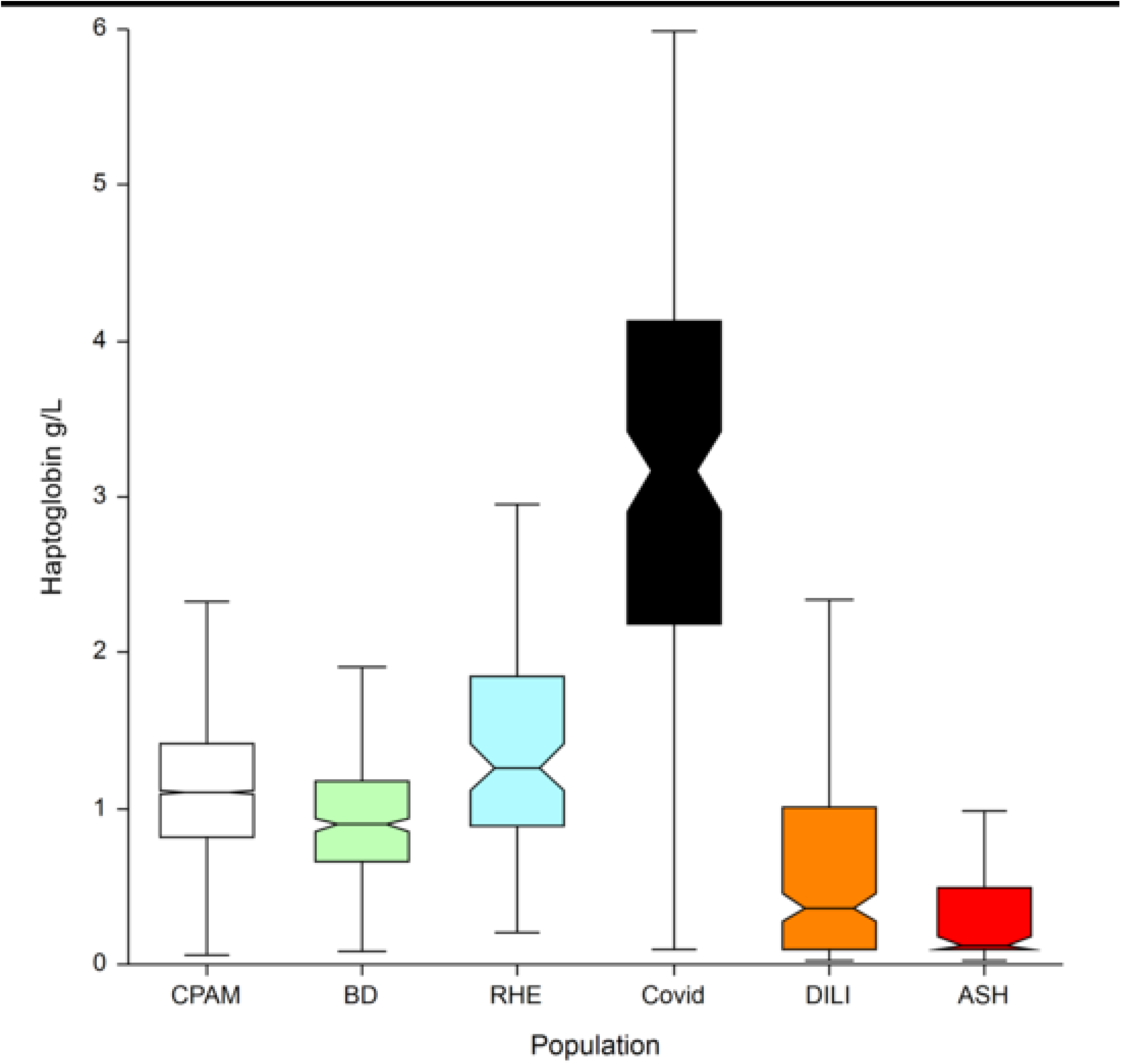
Haptoglobin CPAM: general population ASH: severe acute alcoholic hepatitis, DILI: drug induced liver disease, RHE: rheumatologic disease, BD: blood donors.

The area under the characteristics curve (AUROC;95%CI) in 136 Covid-19 cases and 7,481 controls was 0.978(0.957-0.988), which outperformed haptoglobin and liver function tests (**fig2C**). Apolipoprotein-A1 at a cutoff of 1.25 g/L, had the best Youden index (86.7%) with a sensitivity of 90.6%(84.2-95.1) and a specificity of 96.1%(95.7-96.6) for the diagnosis of Covid-19.

**Fig 2C.**
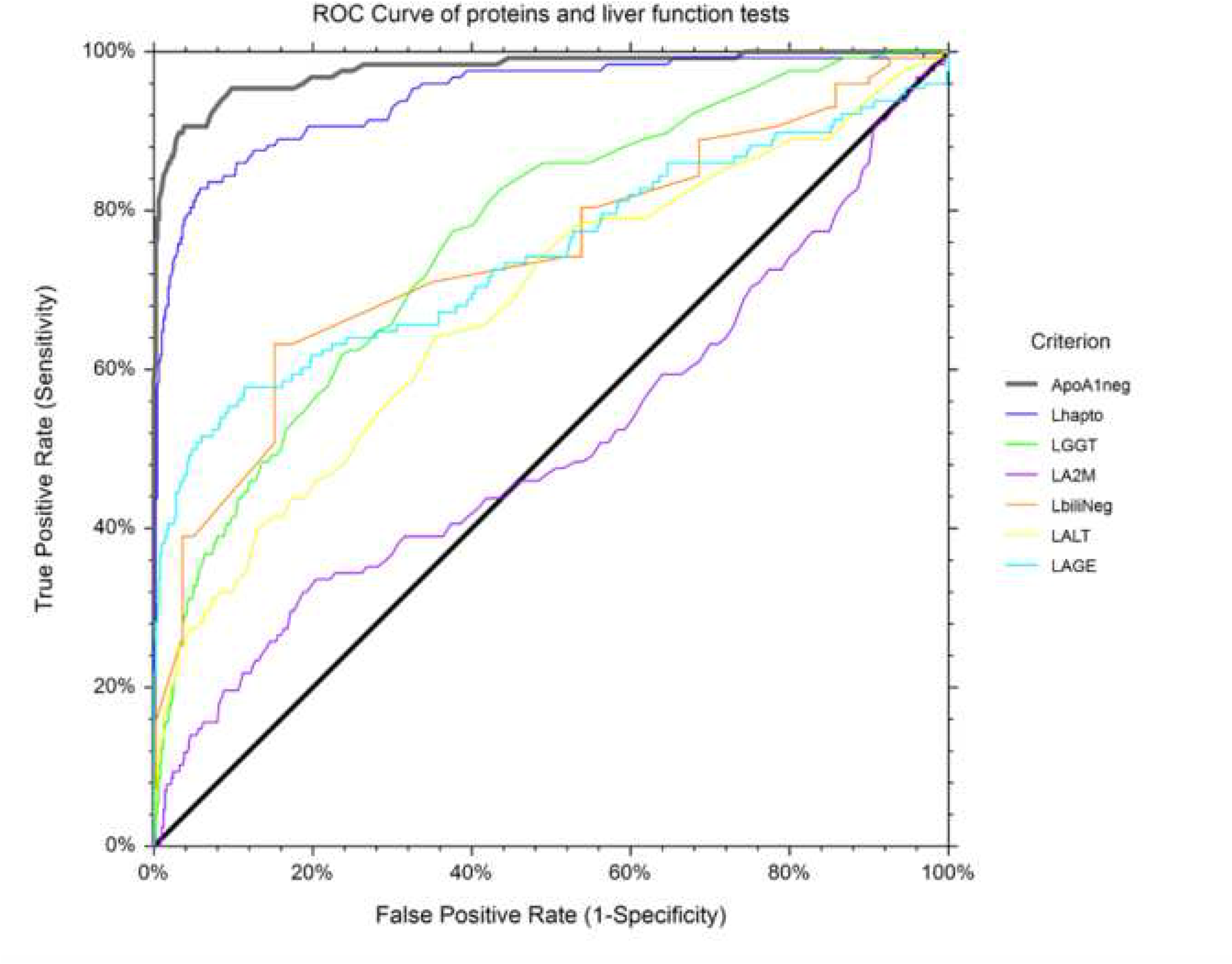
AUROCs of each FibroTest components.

For a prevalence of 1.8% (136/7617;1.5-2.1) of Covid-19 cases, the positive predictive value was 30.0%(25.6-34.7) and the negative predictive value was 99.8%(99.7-99.9). The adjusted predictive values according to prevalence predicted in the French population/^6^ were detailed in **supplementaryFile2** as well as the specificity-sensitivity including blood donors (**supplementaryFig12**), and patients with rheumatological disease (**supplementaryFig13**), and the integrated six databases (**supplementaryFig14**), and the sensitivity in the 19 patients with negative viral nucleic acid testing.

The prognostic value of apolipoprotein-A1 at inclusion for predicting the primary outcome was significant, risk-ratio (RR;95%CI) =5.61 (1.02-31.0;P=0.04), adjusted on age (1.04; 1.01-1.07;P=0.04), GGT (2.88;1.01-8.19;P=0.04). The 71 patients with apolipoprotein-A1 value>=0.84 g/L, the median value at inclusion, had a significant higher survival without ICU (93.0%;87.0-98.9) than the 65 patients with lower value (75.8%; 65.1-86.5; P=0.02) (**fig2D**). Repeated assessments of 305 sera (**supplementaryFig15**) showed that in patients who survived without ICU, the mean apolipoprotein-A1 raised significantly already at 10 days, as well as the 16 patients who survived after their transfer to ICU.

**Fig 2D.**
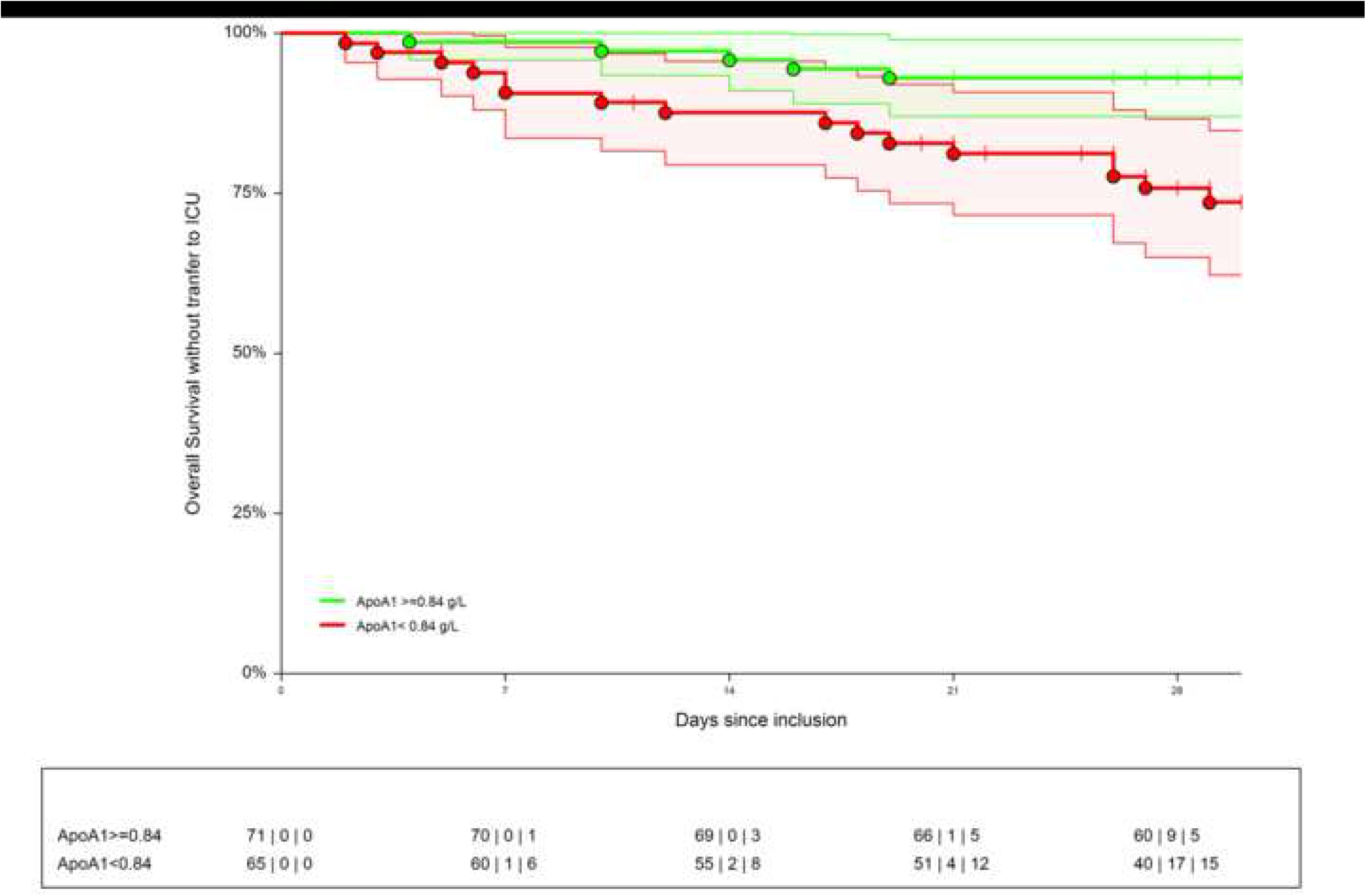
Survival without transfer to intensive care unit (ICU) The 71 patients with apolipoprotein-A1 value >= 0.84 g/L, the median value at inclusion, had a significant higher survival without ICU (93.0%;87.0-98.9) than the 65 patients with lower value (75.8%;65.1-86.5;P=0.02).

The prevalence of diarrhea on initial presentation was 29 out of 131 cases (22.1%;95%CI 15.4-30.2). In this subset, the only significant difference was a lower median number of polynuclear leucocytes (**supplementaryTable5**, **supplementaryFile2**).

## Discussion

Our study shows in three cohorts of patients at risk of liver fibrosis, that apolipoprotein-A1 had a highly significant decrease in 2020 vs previous years, and a highly significant negative daily time-related association with the number of Covid-19 cases. Apolipoprotein-A1 decrease had a high sensitivity in prospective hospitalized patients, with a high specificity in retrospective controls, and an independent prognostic value for the survival without transfer to ICU. These results have certain strengths and limitations.

### Decrease of apolipoprotein-A1 in 2020

The originality of these results was not the decrease in apolipoprotein-A1 during the peak of the pandemic in April, as very low levels of HDL-cholesterol in sera collected in Covid-19 were known in severe pneumonia since 1920 (**supplementaryTable1**).^2^ More intriguing was the very early decrease observed since January 2020 in the USA when the number of Covid-19 cases was unknown. The first known Covid-19 patient was detected on 27/12/2019 and 19/01/2020 in France and the USA, respectively(**supplementaryFile3**). The larger sample size of the US surveillance population, compared to the French, allowed detection of a significant 1% increase in the proportion of subjects possibly infected using the 1.25 g/L cutoff in January (**fig1C**), without any inflammatory signal using haptoglobin. We hypothesized that the SARS-CoV2 virus influenced the liver or intestinal synthesis of apolipoprotein-A1, in asymptomatic patients or in those with unusual mild symptoms.

### Confounding factors

The decrease of apolipoprotein-A1 in 2020 *vs*. previous years, as well as the time-related association of apolipoprotein-A1 in 2020 and Covid-19 might be due to numerous confounding factors. In the context of the pandemic we used cohorts of subjects requiring surveillance of liver fibrosis biomarkers which represent at least 30% of the general adult population in the USA and in France. In these cohorts 70% of the subjects had no or minimal fibrosis. The decrease in apolipoprotein-A1 in 2020 compared to 2019, and 2018 cannot be explained by bias due to gender, age, the cause of liver disease (**table1, supplementaryFile3**), and the severity of liver diseases (**supplementaryFile4**). The prevalence of severe cases cannot explain the significant decrease in apolipoprotein-A1 already observed in January 2020. GGT a very sensitive liver biomarkers did not change during the first 3 months (**supplementaryFig6**). Finally, in these severe liver diseases, haptoglobin should be also significantly decreased (**fig2A**), which was not observed.

In the US-cohort the proportion of sera with NAFLD was increased by 1.8% in 2020 vs 2019 (**table1**). However, after stratification for age and gender, no significant changes were observed for all other biomarkers (**supplementaryFig8**,**supplementaryFig9**,**supplementaryFig10**, **supplementaryFig11**,**supplementaryFile12**). ALT was the only biomarker of the liver tests which increased significantly at the 13 ^th^ week of 2020, above the usual mean value observed in 2019 (**supplementaryFile7**). This increase in ALT was not associated with any other changes (**supplementaryFig10**, **supplementaryFig11**,**supplementaryFig12**). We have no clear explanation. We hypothesize that another confounding factor could be a DILI, including oral acetaminophen or hydroxychloroquine misuse during the pandemic. Such factor could explain an increase in ALT, without increase in haptoglobin, in subjects with mild symptoms.

The absence of haptoglobin change (**supplementaryFig4**) associated with the linear apolipoprotein-A1 decrease (**fig1B**), has never been described before. It was known that in patients with severe fibrosis these two proteins decrease.^10^ It was also known that in severe pneumonia the haptoglobin increase was associated to the apolipoprotein-A1 decrease (**supplementaryTable1**), as we observed in the cohorts with high prevalence of severe Covid-19 (**fig2B**)(**supplementaryFile3**). The 30 weeks followup permitted to see the return to normal values of haptoglobin in these cohorts, associated to the decrease of severe Covid-19 cases admissions (**supplementaryFig4A**). Furthermore, the recovering patients followed by repeated sera in the prospective study, had both a significant increase of apolipoprotein-A1 (**supplementaryFig15**B) and a significant decrease in haptoglobin (**supplementaryFig15**D), 10 days after inclusion.

### Temporal associations between Covid-19 confirmed cases and apolipoprotein-A1 decrease

Our results (**fig1C**) suggest that the spread of the pandemic in the US-cohort would be at least around 5.6%, and at least of 4.3% in the French-cohort. In France, this estimate does not differ from the recent French model that predicted a rate of infection of between 2.8% to 7.2% in the general population.^18^

This temporal association of low apolipoprotein-A1 with the number of confirmed cases, persisted in the two different pandemic changes. In France, after the national lockdown the proportion of low apolipoprotein-A1 returned to the cohort usual 2018-2019 values in June together with the dramatic regression of confirmed cases, still maintained in July. In the US, the apolipoprotein A1 and confirmed cases had the same kinetics with the dramatic increase in April and a plateau in June, still ongoing in July (**fig1E**).

### Specificity of apolipoprotein-A1

There is a high risk of overestimating the specificity of a test in Covid-19 when the participants enrolled in the studies might not be representative of targeted populations.^1^ However, our previous cohorts of patients with severe liver diseases allowed us to identify the major risks of a significant decrease in apolipoprotein-A1 (false positives), mainly due to severe hepatic insufficiency and severe fibrosis. An impact of malnutrition on apolipoprotein-A1 values, was not possible as no changes in weight and height were observed.

### Sensitivity of apolipoprotein-A1

There is also a high risk of overestimating the sensitivity.^1^ It is difficult to confirm the sensitivity for asymptomatic infection, due to the absence of validations of SARS-CoV2-antibodies. To validate the sensitivity of apolipoprotein-A1, a large number of asymptomatic “apparently healthy” subjects who are positive for SARS-CoV-2 viral nucleic acid testing is needed. Our patients included with Covid-19 symptoms had a median of 70 years of age and were severe enough to justify admission to hospital, but none of them required mechanical ventilation at admission, 86% survived, only 6% were transferred to the intensive care unit, and 9% died (**table1**). The sensitivity in the prospective part of our study was similar in the 19 patients who were negative for viral nucleic acid testing (94.7%) to that in positive patients (89.9%). Thus, a simple measurement of apolipoprotein-A1 could be useful for clinicians due to the high percentage of false negatives in available viral nucleic acid testing.^18^Apolipoprotein-A1 or HDL-cholesterol are already being assessed in many large ongoing studies in patients with chronic diseases, which could rapidly validate our results.

### Prognostic performance

The univariate prognostic value of apolipoprotein-A1 was confirmed (**fig2D**) but to our knowledge, it was the first time that its prognostic value persisted after adjustment by age, a marker of acute inflammation (haptoglobin), a sensitive marker of liver injury (GGT), and a marker of liver fibrosis (A2M). Among patients with recovery, there was a significant increase of apolipoprotein-A1 10 days after admission (**supplementaryFig15**). Therefore, apolipoprotein-A1 measurement could help for the decision to transfer to ICU.

### Mechanisms of the early decrease in apolipoprotein-A1 before recognition of the pandemic

We never observed such profile of biomarkers in our experience since 2001 with more than three million of FibroTest assessed in liver diseases (**supplementaryFile1**).^10,12-17^

Although the mechanisms explaining the decrease in apolipoprotein-A1 in late severe Covid-19 pneumonia are known (**supplementaryTable1**), the reason for the early decrease before the acute phase, when haptoglobin remained normal is unclear. In patients with severe pneumonia, apolipoprotein-A1 decrease was associated with acute inflammation and the “cytokine storm” with an increase in IL6 and acute phase proteins such as CRP and haptoglobin. This dissociation suggests that different mechanisms play a role in the early influence of the SARS-CoV2 virus on the synthesis of apolipoprotein-A1, and the intestine could be more involved than the liver. The SARS-CoV2 virus could impact several pathways leading to a decrease in the intestinal synthesis and absorption of apolipoprotein-A1 in the small intestine resulting in the decrease in serum.^3-9^ These mechanisms are discussed in **supplementaryFile1**. The first could the inhibition of lysophosphatidylcholine-acyltransferase-3 activity. There is evidence of direct SARS-CoV2 infection of the endothelial cell and diffuse endothelial inflammation in the intestine.^7,8,9^ SARS-CoV2 uses angiotensin-converting enzyme-2 receptor (ACE-2) expressed on endothelial cells, to infect the host, widely expressed in the lung, intestine and liver.^9,20^ The second mechanism could be an impact of the virus through the intestinal mucus.^21^ Apolipoprotein-A1 is released as a free apolipoprotein from the apical side of enterocytes into the lumen in the fasting state. Apolipoprotein-A1 had faster turnover in mucus, which could be a target for SARS-CoV2.^7,8,21^

### Conclusion

Despite the limitations of this study, these results suggest that apolipoprotein-A1 could be a component of multi-analyte Covid-19 diagnostic and prognostic tests. It could also help to manage patients with a clinical suspicion of Covid-19 and a negative virological test. These results must be validated in independent large cohorts, ideally with virologic and efficiency endpoints. The role of SARS-CoV2 in the possible “asymptomatic” decrease in apoliprotein-A1 could be related to intestinal infection without or before overt pulmonary disease. Finally, one hundred years after surrogate sentinel HDL-cholesterol for pneumonia,^2^ apolipoprotein-A1 for the second time in a century, could be “one of the early warning systems that alert the world to potential outbreaks”.^22^

## Data Availability

The data that support the findings of this study are fully available from the supplementary files.

## Conflict of interest statement: a statement to declare any conflict of interest

Thierry Poynard is the inventor of FibroTest, founder of BioPredictive, the patents belong to the public organization Assistance Publique-Hôpitaux de Paris. Olivier Deckmyn, Valentina Peta, Yen Ngo, Jean Marie Castille, Fabienne Drane and Clemence Franc are full employees of BioPredictive. The other authors have nothing to declare.

## Authors’ contribution

Thierry Poynard, M.D. PhD (Conceptualization: Lead; Data curation: Equal; Formal analysis: Equal; Funding acquisition: Equal; Investigation: Equal; Methodology: Lead; Project administration: Lead; Resources: Equal; Supervision: Lead; Validation: Lead; Visualization: Lead; Writing – original draft: Lead; Writing – review & editing: Lead)

Olivier Deckmyn, PhD (Conceptualization: Equal; Data curation: Equal; Methodology: Equal; Software: Lead; Validation: Equal; Visualization: Equal; Writing – original draft: Equal; Writing – review & editing: Equal)

Marika Rudler, MD, PhD (Data curation: Equal; Investigation: Equal; Resources: Equal)

Valentina Peta, PhD (Data curation: Equal; Investigation: Equal; Resources: Equal; Writing original draft: Equal; Writing review & editing: Equal)

Yen Ngo, MD PhD (Data curation: Equal; Investigation: Equal; Methodology: Equal; Validation: Equal)

Mathieu Vautier, MD (Data curation: Equal; Investigation: Equal; Resources: Equal)

Sepideh Akhavan, MD PhD (Data curation: Equal; Investigation: Equal; Resources: Equal; Validation: Equal)

Vincent Calvez, MD PhD (Resources: Equal; Supervision: Equal; Validation: Equal)

Clemence Franc, MS (Formal analysis: Equal; Investigation: Equal; Writing – original draft: Equal)

Jean Marie Castille, PhD (Funding acquisition: Equal; Project administration: Equal; Validation: Equal)

Fabienne Drane, Ms (Investigation: Equal; Resources: Equal; Validation: Equal)

Mehdi Sakka, MD PhD (Investigation: Equal; Resources: Equal; Validation: Equal)

Dominique Bonnefont-Rousselot, Md PhD (Resources: Equal; Supervision: Equal; Validation: Equal)

Jean Marc Lacorte, MD PhD (Resources: Equal; Supervision: Equal; Validation: Equal)

David Saadoun, MD PhD (Data curation: Equal; Investigation: Equal; Resources: Equal)

Yves Allenbach, MD PhD (Data curation: Equal; Investigation: Equal; Resources: Equal)

Olivier Benveniste, MD PhD (Investigation: Equal; Resources: Equal; Supervision: Equal)

Frederique Gandjbakhch, MD PhD (Data curation: Equal; Investigation: Equal; Resources: Equal)

Julien Mayaux, MD PhD (Data curation: Equal; Investigation: Equal; Resources: Equal)

Olivier Lucidarme, MD PhD (Investigation: Equal; Resources: Equal; Validation: Equal)

Bruno Fautrel, MD PhD (Investigation: Equal; Resources: Equal; Supervision: Equal)

Vlad Ratziu, MD PhD (Formal analysis: Equal; Investigation: Equal; Resources: Equal)

Chantal Housset, MD PhD (Formal analysis: Equal; Supervision: Equal; Validation: Equal; Writing – original draft: Equal)

Dominique Thabut, MD PhD (Project administration: Equal; Resources: Equal; Supervision: Equal; Validation: Equal)

Patrice Cacoub, MD PhD (Conceptualization: Lead; Formal analysis: Lead; Funding acquisition: Equal; Investigation: Equal; Project administration: Lead; Resources: Equal; Supervision: Lead; Validation: Lead; Visualization: Lead; Writing – original draft: Equal; Writing review & editing: Equal)

